# On the Environmental Determinants of COVID-19 Seasonality

**DOI:** 10.1101/2021.03.01.21252243

**Authors:** Yeon-Woo Choi, Alexandre Tuel, Elfatih A. B. Eltahir

## Abstract

Viral respiratory diseases (VRDs), such as influenza and COVID-19, are thought to spread faster over winter than during summer. It has been previously argued that cold and dry conditions were more conducive to the transmission of VRD than warm and humid climates, although this relationship appears restricted to temperate regions, and the causal relationship is not well understood. The severe acute respiratory syndrome coronavirus 2 (SARS-CoV-2) causing COVID-19 has emerged as a serious global public health problem after the first COVID-19 reports in Wuhan, China, in late 2019. It is still unclear whether this novel respiratory disease will ultimately prove to be a seasonal endemic disease. Here, we suggest that Air Drying Capacity (ADC; an atmospheric state-variable known to control the fate/evolution of the virus-laden droplets) and ultraviolet radiation (UV) are probable environmental determinants in shaping the transmission of COVID-19 at the seasonal time scale. These variables, unlike temperature and humidity, provide a physically-based framework consistent with the apparent seasonal variability in COVID-19 prevalence across a broad range of climates (e.g., Germany and India). Since this disease is known to be influenced by the compounding effect of social, biological, and environmental determinants, this study does not claim that these environmental determinants exclusively shape the seasonality of COVID-19. However, we argue that ADC and UV play a significant role in COVID-19 dynamics at the seasonal scale. These findings could help guide the development of a sound adaptation strategy against the pandemic over the coming seasons.

**Plain Language Summary:** There is growing scientific interest in the potential seasonality of COVID-19 and its links to climate variables. This study aims to determine whether four environmental variables, namely temperature, humidity, Air Drying Capacity (ADC), and ultraviolet radiation (UV), are probable environmental determinants for the observed seasonal dynamics of COVID-19 prevalence, based on extensive country-level data spanning the first year of the pandemic. Although the influence of socio-economic factors may be dominant, we here suggest that ADC and UV are key environmental determinants of COVID-19 and can potentially affect the transmission and seasonality of the disease across a wide range of climates.

**Key Points:**

- The seasonality of COVID-19 appears to follow seasonality of some environmental variables.
- Seasonality of air drying capacity and ultraviolet radiation consistently match seasonality of COVID-19, across climatic zones.
- Seasonality of air humidity and temperature, match seasonality of COVID-19 in temperate climates, but not in tropical monsoon climates.

## 1 Introduction

Since the 2019 novel coronavirus responsible for COVID-19 was initially reported in December 2019 in Wuhan, the epicenter of the current pandemic (Li et al., 2020; Zhou et al., 2020), there has been growing scientific interest in the seasonality of this novel disease and the potential influence of climate variables. However, the analysis on this issue has been complicated by limited data. Nonetheless, based on epidemiological studies involving other Viral Respiratory Diseases (VRDs; e.g., influenza), which, like COVID-19, are mainly transmitted by contact, droplets, and fomites (Stilianakis and Drossinos, 2010; Dhand and Li, 2020), it may be possible to provisionally infer not only the seasonal nature of COVID-19, but also the role of environmental factors in shaping this seasonality.

A well-known and well-studied VRD is influenza, which tends to peak during winter in temperate regions (e.g., Shaman and Khon, 2009; Tamerius et al., 2011; 2013; Ballester et al., 2016; Choi et al., 2020). Previous studies have attempted to understand its seasonality by considering the effects of environmental/weather conditions on virus survival and transmissibility. Based on experiments with inoculated guinea pigs, Lowen et al. (2007) reported that the transmission of the influenza virus was significantly suppressed under high absolute humidity and warm temperature conditions. Subsequent research based on similar laboratory experiments emphasized the role of absolute humidity over relative humidity as a key environmental factor of influenza transmission (Shaman and Kohn, 2009). The analysis of data from temperate regions further supported the hypothesis that low specific humidity conditions favoured the survival and transmission of the influenza virus at the scale of populations, and could be the main cause of winter epidemics (Shaman et al., 2010; 2011).

However, this perspective failed to account for influenza dynamics in tropical countries, where the disease typically peaks during the wet season (Tamerius et al., 2011). Further research argued that seasonality in tropical regions could be explained by a modulation of the effect of absolute humidity with temperature, with “cold-dry” and “warm-wet” conditions favoring influenza transmission (Tamerius et al., 2013; Deyle et al., 2016). Yet, it is still unclear why influenza would respond differently to absolute humidity at different temperatures, and why different statistical relationships would be needed for countries of different climates and latitudes. In addition, the exact mechanisms by which absolute humidity may affect the survival and transmission of the influenza virus remain unknown. Much of the attention has also focused on whether the environment affected virus survival and host contagiousness. Yet, since influenza and VRDs in general are transmitted in large part through respiratory droplets, environmental conditions may also affect VRD prevalence through their effect on the fate of these droplets. In an earlier study, we proposed by contrast that the prevalence of influenza, and VRDs in general, was likely to be shaped instead by the Air Drying Capacity (ADC; see Section 2.2), a state variable involving both temperature and humidity, which controls the evolution of respiratory droplets and is based on Maxwell’s theory of droplet evolution via coupled heat and mass transfer (Maxwell, 2003; Choi et al., 2020).

Regarding COVID-19, most of the virus prevalence during the initial stage of the pandemic was found in areas with specific climate conditions: average temperatures of 5-11 °C, combined with low specific humidity of 3-6 g/kg (Sajadi et al., 2020); and temperature range of 3 to 17 °C with humidity between 4 and 9 g/m^3^ (Bukhari and Jameel, 2020). Further studies also argued that low temperature and low humidity were major drivers behind the explosive increase in confirmed COVID-19 cases, because they weakened the host’s immune system and increased virus stability and transmission (e.g., Araújo and Naimi, 2020; Wang et al., 2020a; Wu et al., 2020; Chin et al., 2020; Moriyama et al., 2020). On the other hands, several studies have emphasized the effect of atmospheric humidity on the prevalence of this disease, consistent with influenza dynamics (Baker et al., 2020; Ward et al., 2020; Ma et al., 2020). By contrast, the spread of COVID-19 was found to be more correlated with temperature than humidity in certain areas (Bherwani et al., 2020; Kaplin et al., 2020; Tosepu et al., 2020; Xie and Zhu, 2020). In addition, attention has also focused on Ultraviolet (UV) radiation as a potential environmental factor impacting VRD transmission (Carleton et al., 2021). Laboratory and epidemiological studies have shown that strong UV radiation appeared to have a profound effect on the survival of SARS-Cov-2 and other coronaviruses (Duan et al., 2003; Darnell et al., 2004; Seyer and Sanlidag 2020; Schuit et al., 2020; Ratnesar-Shumate et al., 2020).

These studies provide some evidence that environmental variables play some role on COVID-19 transmission, even though it is generally agreed that many determinants, including social, biological, and environmental factors, shape the transmission of VRDs. In particular, the massive measures taken by governments in response to the COVID-19 pandemic have strongly impacted its dynamics (Bherwani et al., 2020; Wang et al., 2020a). Additionally, most studies have focused on the initial months of the pandemic, when data was too limited to detect strong seasonal signals (Sajadi et al., 2020; Bukhari and Jameel, 2020; wang et al., 2020a). Therefore, at this stage, it remains unclear to what extent COVID-19 prevalence exhibits seasonality, whether this seasonality is shaped by environmental factors, and if so, which are the most important. Here, we focus on these questions by examining relationships between COVID-19 prevalence and environmental variables across a wide range of climates, based on available data covering the first year of the pandemic.

## 2 Data and Method

### 2.1 Datasets

6-hourly temperature, dew point temperature and surface pressure data, and hourly UV fields are taken from the ERA5 reanalysis (Hersbach et al. 2018) at 0.25° × 0.25° horizontal resolution, for the period ranging from March 1^st^, 2020 to January 13^th^, 2021. With this data we calculate daily mean temperature, specific humidity, and ADC (see Section 2.2). Since population is not uniformly distributed within each country, we avoid simple country-averaged climate variables which do not correctly reflect the climate conditions to which the population is exposed. Instead, we calculate population-weighted average temperature, specific humidity, ADC, and UV across all grid cells contained in a given country, based on weights obtained from a gridded distribution of population density, following Carleton et al. (2021). Population density data is taken from the gridded population of the world (GPW) v4 dataset (CIESIN, 2018).

Daily data on confirmed COVID-19 cases, number of tests, stringency index (i.e., a composite index of political measures) and population for each country are from the “Our World in Data” database (available at https://ourworldindata.org/). Subnational-level COVID-19 epidemiological data for the Australia, China, and Canada are available at the Johns Hopkins University Center for Systems Science and Engineering (JHU CCSE; https://data.humdata.org/). Daily COVID-19 data at the scale of different states within the United States are provided at the COVID Tracking Project (available at https://covidtracking.com/). A threshold of at least 10,000 cumulative COVID-19 tests per 1 million people was retained to discard countries with unrepresentative data. This criterion can somewhat ensure the reliability of the data, although it still has severe limitations (see discussion). This approach yields 54 countries in the temperate Northern Hemisphere, and six tropical countries (Table 1), which are predominantly influenced by tropical monsoon systems with hot-humid summers. To isolate the role of environmental factors in modulating the spread and potential seasonality of COVID-19, five representative countries are carefully selected, which have different climate conditions and constant social controls (i.e., stringency index does not change significantly) over the analysis period (Fig. S1). The list of analyzed countries are provided in Table 1. To consider the incubation period of COVID-19 which is generally regarded as 4 to 7 days but can be as long as 10 days and uncommonly even longer, we mainly use biweekly average or biweekly cumulative values (Li et al., 2020).

**Table 1.** List of countries used in this study.

|  | **List of countries** |
| --- | --- |
| Five representative countries of different climates and latitudes | Canada, Germany, India, Ethiopia, and Chile |
| 54 temperate countries in the Northern Hemisphere | Andorra, Austria, Bahrain, Belarus, Belgium, Bosnia and Herzegovina, Bulgaria, Croatia, Cyprus, Denmark, Estonia, Finland, Germany, Greece, Hungary, Iceland, Iran, Iraq, Ireland, Israel, Italy, Japan, Jordan, Kazakhstan, South Korea, Latvia, Lithuania, Luxembourg, Malta, Mongolia, Morocco, Nepal, Netherlands, North Macedonia, Norway, Pakistan, Poland, Portugal, Qatar, Romania, Russia, Serbia, Slovakia, Slovenia, Spain, Switzerland, Tunisia, Turkey, Ukraine, United Arab Emirates, United Kingdom, Canada, China, US |
| 6 tropical monsoon countries | Bangladesh, Ethiopia, Thailand, Ghana, India, Cote d'Ivoire |

### 2.2 Air Drying Capacity (ADC)

The current mechanistic understanding of VRD transmission assumes that VRDs are mainly transmitted through virus-laden droplets between infectious and susceptible individuals (Wells, 1934; Xie et al.,2007; Bourouiba, 2020). Since droplets tend to evaporate more quickly the higher the temperature and the lower the humidity (Wang et al., 2020b), these two variables could theoretically affect the spread of the VRD by controlling the fate of droplets in their surrounding environment (Maxwell, 2003; Choi et al., 2020). In our earlier study, we proposed a physically-based atmospheric state-variable, Air Drying Capacity (ADC), as a relevant environmental determinant for the transmission of VRD (Choi et al., 2020). We define ADC, (in mm^2^/hr) as the rate of decrease of the droplet surface area, under given temperature and humidity conditions. ADC is an integrated measure of temperature and humidity and is nonlinearly proportional to temperature but inversely proportional to humidity (Table 2). ADC is defined as:

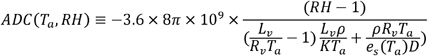

**Table 2.** Relationship between biweekly environmental variables including temperature (deg C), specific humidity (g/kg), ADC (mm^2^/hr), and UV (W/m^2^) for the period from March 1^st^ 2020 to January 13^th^ 2021 across countries in the temperate Northern Hemisphere. The values in parentheses is same as those without parentheses, but for tropical monsoon countries.

|  | Temperature | Specific humidity | ADC | UV |
| --- | --- | --- | --- | --- |
| Temperature | 1 (1) | 0.7 (0.7) | 0.6 (0.0) | 0.6 (0.0) |
| Specific humidity |  | 1 (1) | 0.2 (0.3) | 0.3 (0.0) |
| ADC |  |  | 1 (1) | 0.6 (0.5) |
| UV |  |  |  | 1 (1) |

where *T*_*a*_ is the ambient temperature,*RH* the ambient relative humidity, *L*_*v*_ the latent heat of vaporisation, *R*_*v*_ the specific gas constant for water vapour, *ρ* liquid water density, *K* the thermal conductivity of air, *e*_*s*_ the saturation vapor pressure at temperature, *D* is the diffusion coefficient of water vapour. More details can be found in Choi et al. (2020).

## 3 Results

The prevalence of COVID-19 tends to exhibit a distinct seasonality in the five representative countries (see Section 2.1 for a selection of countries), similar to that of influenza (Figs. 1 and S2; Tamerius et al., 2011; 2013; Choi et al., 2020). For example, temperate countries in both hemispheres (i.e., Canada, Germany, and Chile) experienced peak COVID-19 incidence in their respective winter months. These regions in temperate climates exhibited over the past year opposite evolutions of COVID-19 and of the four environmental variables we consider (temperature, specific humidity, ADC, and UV). That is, low values in temperature, humidity, ADC, and UV generally occurred in the months of highest COVID-19 incidence.

**Figure 1.**
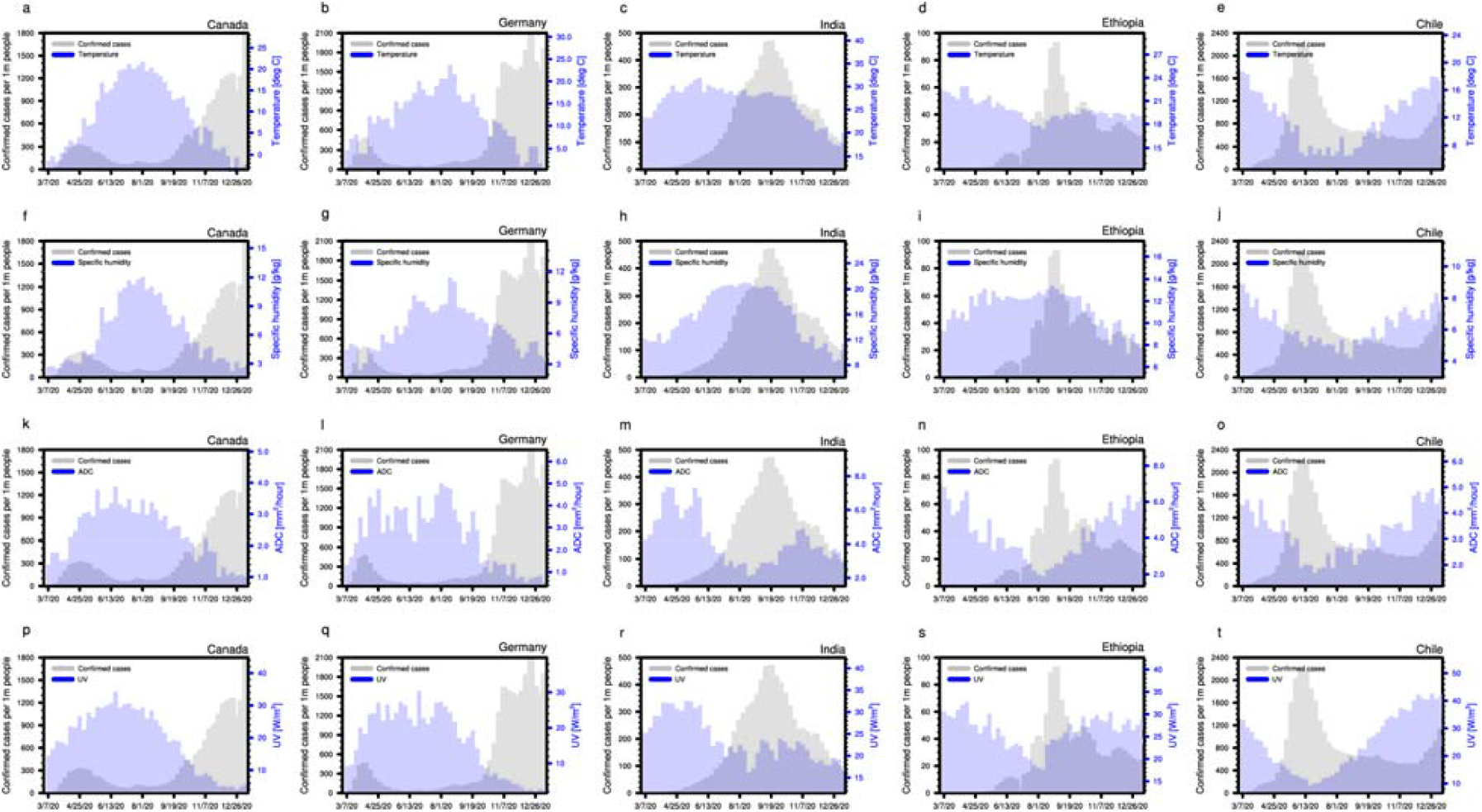
Seasonal variation of COVID-19 prevalence and environmental variables. Seasonal variation of weekly COVID-19 prevalence alongside weekly (a-e) temperature, (f-j) specific humidity, (k-o) ADC, and (p-t) UV across COVID-19 hotspots, such as (a, f, k, and p) Canada in North America, (b, g, l, and q) Germany in Europe, (c, h, m, and r) India in Asia, (d, i, n, and s) Ethiopia in Africa, and (e, j, o, and t) Chile in South America.

On the other hand, COVID-19 appears to have peaked during northern hemisphere summer in tropical regions (i.e., India, and Ethiopia), specifically at the time of the monsoons when specific humidity was at its highest. There, the seasonality of COVID-19 and that of temperature and specific humidity thus turn out to be inconsistent with what is observed for temperate regions, as is the case for influenza (Tamerius et al., 2011). The seasonal evolution of ADC and UV, by contrast, is consistent with COVID-19 prevalence in all countries (Figs 1k to 1t). High UV and high ADC linked to lower prevalence, and vice-versa, although this relationship is weak over a certain period in autumn over India. This result is in line with the expected effect of these two variables on the survival and transmissibility of SARS-Cov-2 (e.g., Choi et al., 2020; Seyer and Sanlidag, 2020; Schuit et al., 2020; Ratnesar-Shumate et al., 2020). The case of India in particular is quite striking, especially since the country attracted much attention due to the explosiveness of its COVID-19 outbreak and its different timing of peak incidence (e.g., Bherwani et al., 2020) compared to those in temperate regions.

Pooling the data for the five representative countries together and conditioning biweekly new COVID-19 cases on the various environmental variables yields further insights consistent with the previous analysis (Fig. 2). While COVID-19 spread appears lowest at high temperatures, the peak in COVID-19 spread occurs at somewhat average temperatures and the relationship is altogether inconsistent. Similarly, new cases are at their highest at low specific humidity values, but almost as high when specific humidity is large as well. By contrast, the conditioning on ADC and UV stands out as much more consistent: (1) COVID-19 prevalence nonlinearly increases as ADC approaches zero, in keeping with its effect on virus-laden droplets exhaled by infected patients (Choi et al., 2020); (2) similarly, low UV values appear to provide favorable conditions for the spread of the disease, consistent with the impact of UV light on the survival of SARS-CoV-2 (Seyer and Sanlidag, 2020; Schuit et al., 2020; Carleton et al., 2021); and (3) the lowest ADC (0-2 mm^2^/hr) and UV (2-10 W/m^2^) values with a probability less than 15% are systematically linked to the largest numbers of COVID-19 infections (Figs S2, 2c, 2d, 2g, and 2h). Given the relatively similar and constant level of social control in the five analyzed countries, these findings suggest that ADC and UV, unlike temperature and humidity, are likely environmental determinants of COVID-19 spread and its seasonality.

**Figure 2.**
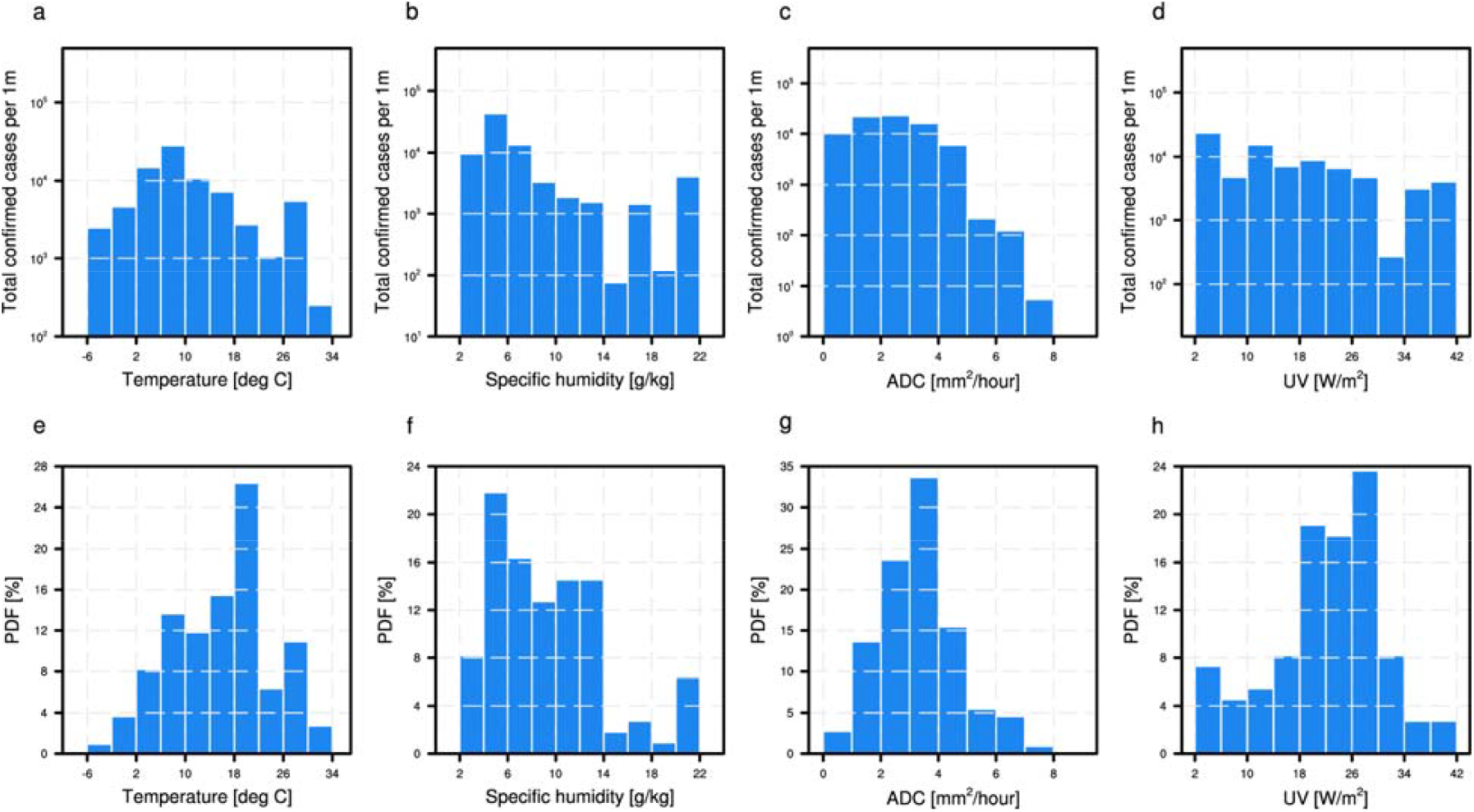
Biweekly environmental variables and biweekly COVID-19 prevalence. Total number of COVID-19 cases across the five representative countries (i.e., Canada, Germany, India, Ethiopia, and Chile) for the period from March 1^st^, 2020 to January 13^th^, 2021 as a function of (a) temperature, (b) specific humidity, (c) ADC, and (d) UV. Probability density function of (e) temperature, (f) specific humidity, (g) ADC, and (h) UV over the region for the same period.

To further advance our understanding of the possible link between COVID-19 seasonality and the four environmental variables, we now analyze data from all available countries with sufficient number of tests, divided into temperate countries in the Northern Hemisphere, and tropical, monsoon-dominated countries (see Section 2.1; Table 1). We find supportive evidence for strong negative relationships between all four environmental variables and COVID-19 spread in the temperate group (Figs 3a-3d, and 4a-4d). Interestingly, in the early stages of pandemic (i.e., first half of the year), ADC and UV exhibit more consistent variability with COVID-19 infections rather than temperature and humidity (i.e., COVID-19 prevalence, ADC, and UV does not change very much, while temperature and humidity increase sharply; Figs. 3a-3d). The discrepancy among them is more obvious in tropical monsoon regions, as expected from results shown in Figs 1-2, and previous influenza studies (e.g., Tamerius et al., 2011). Consistent with the previous 5-country analysis, the tropical group, with a sharp rise in new COVID-19 cases during the wet season, stands in contrast to temperate group. Only ADC and UV, unlike temperature and humidity, show seasonal variations consistent with those of COVID-19 spread across the two groups of countries (Figs. 3g-3h).

**Figure 3.**
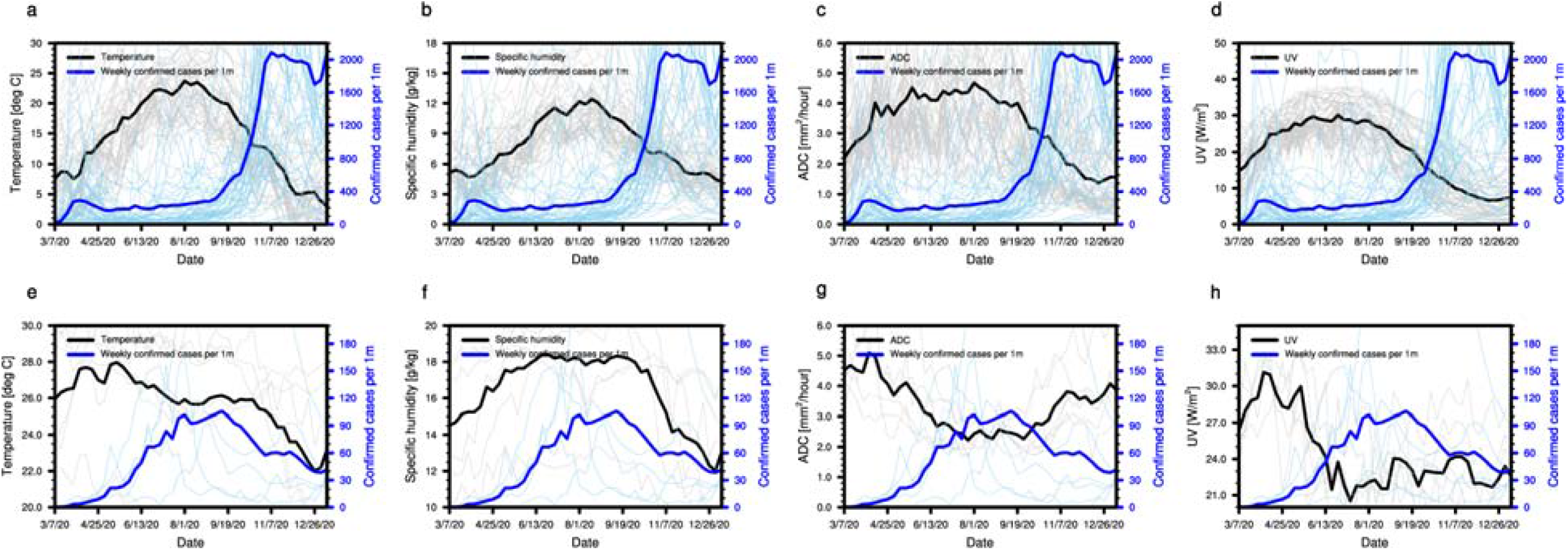
Seasonal variation of COVID-19 prevalence and environmental variables. Seasonal variation of weekly COVID-19 prevalence alongside weekly (a, e) temperature, (b, f) specific humidity, (c, g) ADC, and (d, h) UV across (a-d) 54 temperate countries in the Northern Hemisphere (Table 1) and (e-h) 6 tropical monsoon countries (Table 1).

Conditioning once more COVID-19 cases on the values of the four environmental variables, different statistical relationships are found for the two selected climate zones. The temperate Northern Hemisphere is characterized by a nonlinear decrease of COVID-19 cases with all four variables (Fig. 4), whereas in tropical monsoon regions, COVID-19 cases increase with both temperature and humidity (Fig. 5). By contrast, ADC and UV both show consistent negative relationships with COVID-19 regardless of the region (Figs 4-5). The spread of COVID-19 seems in particular to be very constrained at ADCs of 5 mm^2^/hr or larger. Its sensitivity is also obviously nonlinear, consistent with Choi et al. (2020). Additionally, low UV values (0 to 15 W/m^2^), though quite rare at the annual scale, systematically occur in conjunction with high numbers of new COVID-19 cases, especially in the temperate regions (Figs. 3d, and 4d).

**Figure 4.**
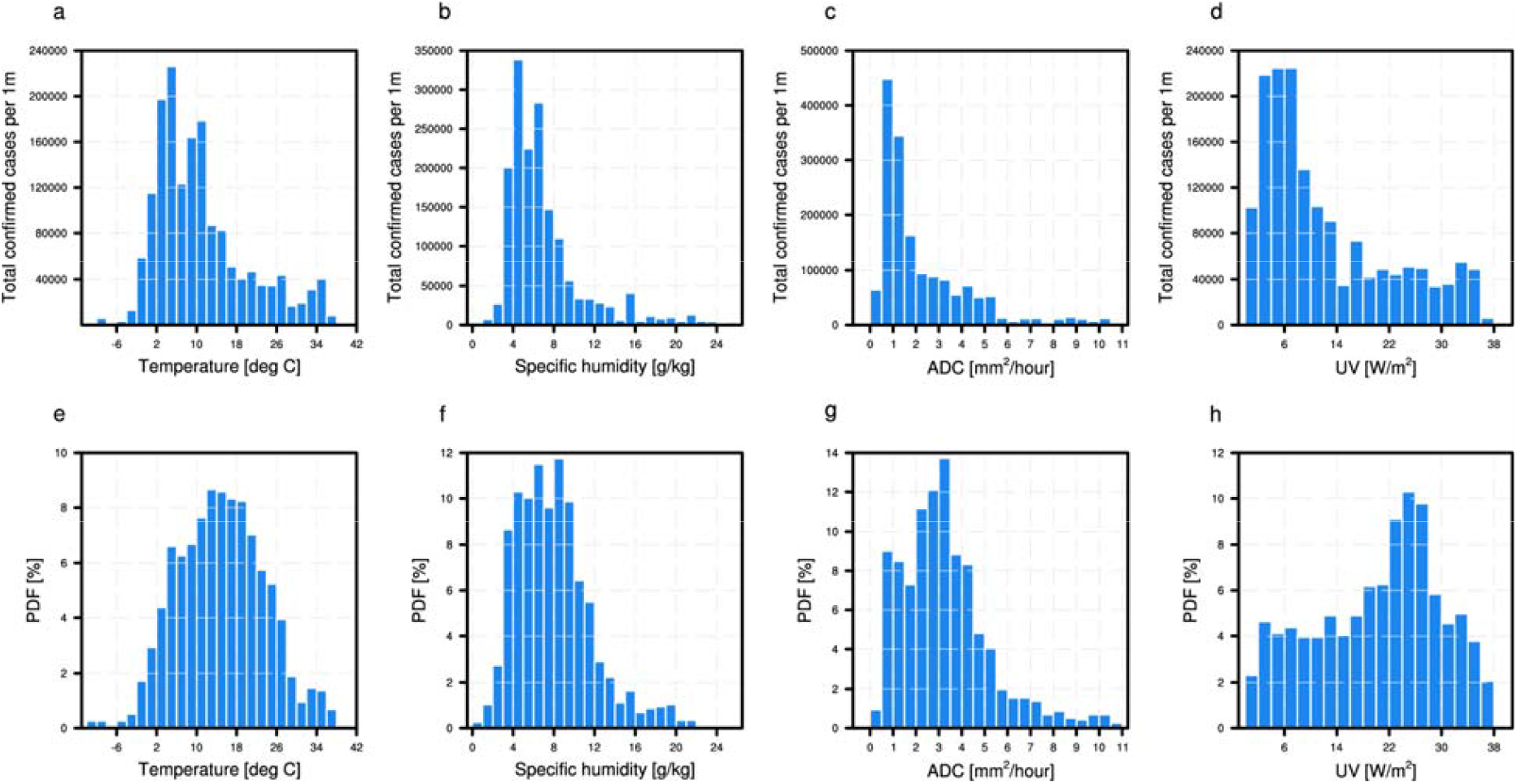
Biweekly environmental variables and biweekly COVID-19 prevalence. Total number of COVID-19 cases across the 54 temperate countries (Table 1) in the Northern Hemisphere for the period from March 1^st^, 2020 to January 13^th^, 2021 as a function of (a) temperature, (b) specific humidity, (c) ADC, and (d) UV. Probability density function of (e) temperature, (f) specific humidity, (g) ADC, and (h) UV over the region for the same period.

**Figure 5.**
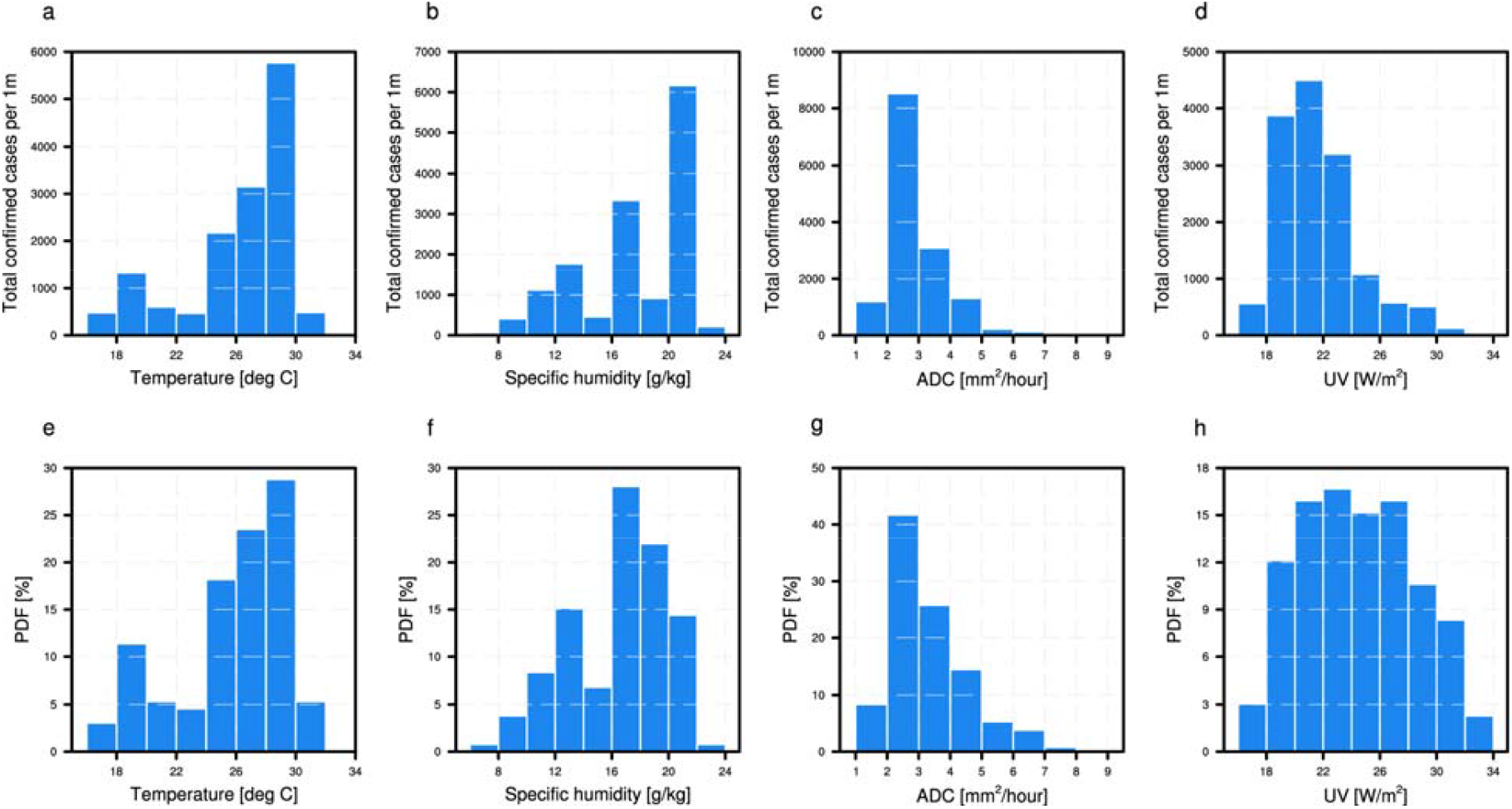
Biweekly environmental variables and biweekly COVID-19 prevalence. Total number of COVID-19 cases across the 6 tropical monsoon countries (Table 1) for the period from March 1^st^, 2020 to January 13^th^, 2021 as a function of (a) temperature, (b) specific humidity, (c) ADC, and (d) UV. Probability density function of (e) temperature, (f) specific humidity, (g) ADC, and (h) UV over the region for the same period.

## 4 Discussion and conclusion

Through extensive country-level data spanning the 1-year pandemic period, this study evaluates and compares the role of four environmental variables (temperature, humidity, ADC, and UV) with the potential to influence the spread of COVID-19. Three important results that emerge from this study are that: (1) the spread and prevalence of COVID-19 appears to display some seasonality consistent, to some extent, with the seasonality of the analyzed environmental variables and their known relationships to influenza seasonality; (2) two environmental variables in particular, ADC and UV, evolve consistently with COVID-19 spread across the selected countries of all climates and at both the weekly and the seasonal timescales, and their relationship to COVID-19 spread is consistent with their respective effects on the fate of respiratory droplets (ADC, Choi et al. 2020) and on virus survival (UV, Seyer and Sanlidag, 2020; Schuit et al., 2020; Ratnesar-Shumate et al., 2020). Therefore, ADC and UV may provide the basis for a physically-based framework shaping seasonal variations in COVID-19 prevalence across the world; and (3) we also find that, as was the case for influenza, absolute humidity and temperature do not exhibit a consistent relationship to COVID-19 cases across all climate zones. Their statistical relationships change directions whether one considers temperate or tropical monsoon countries, which casts doubt as to their true relevance for COVID-19 spread at the country scale, all the more so as the physical mechanisms that could explain these relationships and why they differ depending on the background climate remain unknown. By contrast, the statistical relationship between COVID-19 prevalence and ADC or UV is both consistent across climate zones and with the known physical effects of these two variables on disease transmission. Naturally, all four variables are, to some extent, correlated with each other (Table 2). Temperature and humidity tend to evolve in concert, as warmer air generally holds more moisture. Similarly, for temperate countries, both ADC and UV tend to reach their minimum during winter, when both temperature and specific humidity are also lowest. But such is not the case in tropical, monsoon-dominated countries.

Before concluding, it is important to mention that this study naturally presents several important caveats. First, as for any COVID-19-related study, the quality, extensiveness and uniformity of the data is subject to caution. COVID-19 case data is strongly impacted by testing rates and policies, which differ in space (between different countries) but also in time, since testing protocols have evolved over time. Although this study applied the threshold of at least 10,000 cumulative COVID-19 tests per 1 million people to discard countries with unrepresentative data, but considering all countries without quality control may provide other conclusions that are inherent with high uncertainty. Influenza data also suffers from the same biases, which are inevitable in any global study. Additionally, the spread of COVID-19 is largely shaped by policy measures, such as social distancing, mask mandates, school closures, and event cancellations (Bherwani et al., 2020; Poirier et al., 2020). Finally, we did not consider the effects of indoor environments (e.g., heating and air conditioning), which may also influence the seasonality of VRD (e.g., Xie et al., 2007; Shaman and Kohn, 2009). As always in such cases, one must be careful in concluding about the role of environmental variables in shaping VRD dynamics (Carlson et al., 2020; Zaitchik et al., 2020). Even though vaccines have recently been developed and are currently being administered, and though it is hoped that SARS-Cov-2 will soon be controlled effectively, there is still a possibility that this virus will stay with us as a seasonal disease with milder effects, like influenza. Our findings could therefore help guide the development of a sound adaptation strategy against COVID-19 over the coming years.

## Data Availability

6-hourly temperature, dew point temperature and surface pressure data, and hourly UV fields are taken from the ERA5 reanalysis (Hersbach et al. 2018) at 0.25 by 0.25 horizontal resolution, for the period ranging from March 1st, 2020 to January 13th, 2021. With this data we calculate daily mean temperature, specific humidity, and ADC (see Section 2.2). Since population is not uniformly distributed within each country, we avoid simple country-averaged climate variables which do not correctly reflect the climate conditions to which the population is exposed. Instead, we calculate population-weighted average temperature, specific humidity, ADC, and UV across all grid cells contained in a given country, based on weights obtained from a gridded distribution of population density, following Carleton et al. (2021). Population density data is taken from the gridded population of the world (GPW) v4 dataset (CIESIN, 2018).
Daily data on confirmed COVID-19 cases, number of tests, stringency index (i.e., a composite index of political measures) and population for each country are from the Our World in Data database (available at https://ourworldindata.org/). Subnational-level COVID-19 epidemiological data for the Australia, China, and Canada are available at the Johns Hopkins University Center for Systems Science and Engineering (JHU CCSE; https://data.humdata.org/). Daily COVID-19 data at the scale of different states within the United States are provided at the COVID Tracking Project (available at https://covidtracking.com/). A threshold of at least 10,000 cumulative COVID-19 tests per 1 million people was retained to discard countries with unrepresentative data. This criterion can somewhat ensure the reliability of the data, although it still has severe limitations (see discussion). This approach yields 54 countries in the temperate Northern Hemisphere, and six tropical countries (Table 1), which are predominantly influenced by tropical monsoon systems with hot-humid summers. To isolate the role of environmental factors in modulating the spread and potential seasonality of COVID-19, five representative countries are carefully selected, which have different climate conditions and constant social controls (i.e., stringency index does not change significantly) over the analysis period (Fig. S1). The list of analyzed countries are provided in Table 1. To consider the incubation period of COVID-19 which is generally regarded as 4 to 7 days but can be as long as 10 days and uncommonly even longer, we mainly use biweekly average or biweekly cumulative values (Li et al., 2020).

https://data.humdata.org/

https://covidtracking.com/

## Acknowledgments

We acknowledge support from the Breene M Kerr chair at MIT.

## Author Contributions

E. A. B. E. devised and supervised the study. Y. C. and A. T. carried out analyses. All authors contributed to the manuscript.

## Correspondence

Correspondence and requests for materials should be addressed to

