## Supplementary Material for "On the Environmental Determinants of COVID-19 Seasonality"

21th February, 2020

[1]Ralph M. Parsons Laboratory, Massachusetts Institute of Technology, Cambridge, Massachusetts 02139, USA

†Current affiliation: Institute of Geography, Oeschger Centre for Climate Change Research, University of Bern, Bern, Switzerland.

### Contents

- Figure S1
- Figure S2

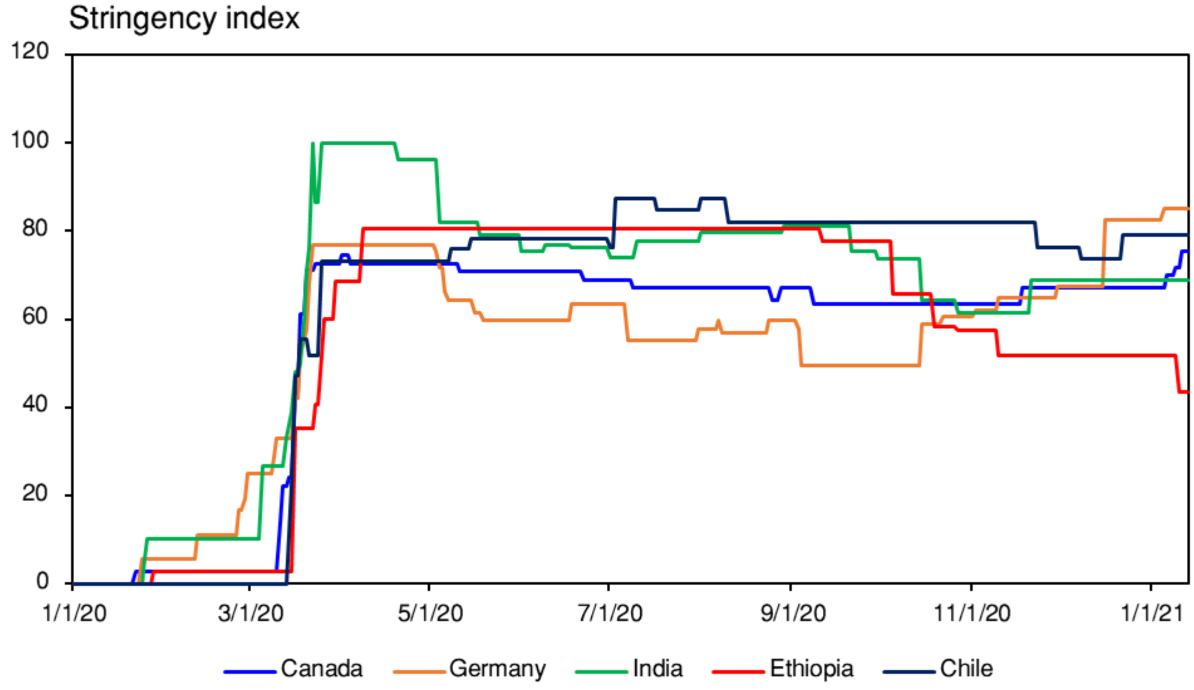


Figure S1: Government Stringency Index. Time series of government stringency index for five representative countries (Table 1). This index is a composite measure based on nine response indicators including school closures, workplace closures, and travel bans.

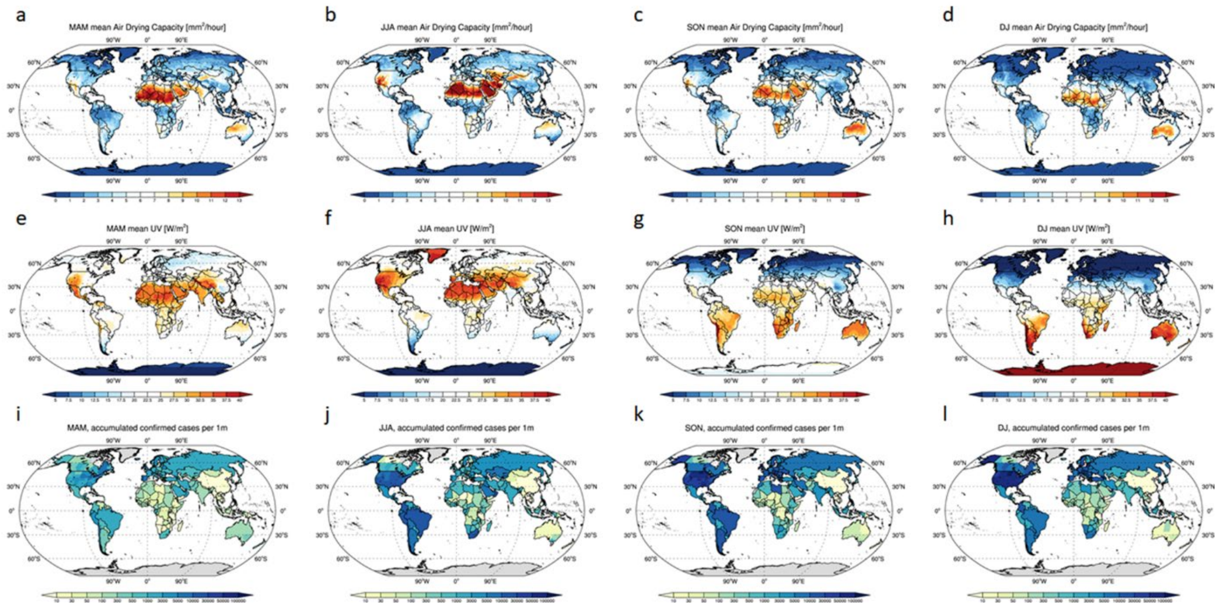


Figure S2: Seasonal mean COVID-19 prevalence and environmental variables. Spatial distribution of seasonal mean (a-d) ADC, (e-h) UV, (i-l) total confirmed COVID-19 cases per 1 million people for (a, e, and i) spring (March to May), (b, f, and j) summer (June to August), (c, g, and k) autumn (September to November), and (d, h, and l) winter (December to January).
